# Preliminary optimisation of simplified sample preparation method to permit direct detection of SARS-CoV-2 within saliva samples using reverse-transcription loop-mediated isothermal amplification (RT-LAMP)

**DOI:** 10.1101/2020.07.16.20155168

**Authors:** Emma L. A. Howson, Stephen P. Kidd, Jason Sawyer, Claire Cassar, David Cross, Tom Lewis, Jess Hockey, Samantha Rivers, Saira Cawthraw, Ashley Banyard, Michael Andreou, Nick Morant, Duncan Clark, Charlotte Walsh, Shailen Laxman, Rebecca Houghton, Joanne Slater-Jefferies, Paula Costello, Ian Brown, Nicholas Cortes, Keith M. Godfrey, Veronica L. Fowler

**Author notes:** Joint first authors (Emma L. A. Howson, Stephen P. Kidd).

## Abstract

We describe the optimization of a simplified sample preparation method which permits rapid and direct detection of SARS-CoV-2 RNA within saliva using reverse-transcription loop-mediated isothermal amplification (RT-LAMP). Treatment of saliva samples prior to RT-LAMP by dilution 1:1 in Mucolyse™, followed by dilution (within the range of 1:5 to 1:40) in 10% (w/v) Chelex^©^ 100 Resin and a 98°C heat step for 2 minutes enabled detection of SARS-CoV-2 RNA in all positive saliva samples tested, with no amplification detected in pooled negative saliva. The time to positivity for which SARS-CoV-2 RNA was detected in these positive saliva samples was proportional to the real-time reverse-transcriptase PCR cycle threshold (C_T_), with SARS-CoV-2 RNA detected in as little as 05:43 (C_T_ 21.08), 07:59 (C_T_ 24.47) and 08:35 (C_T_ 25.27) minutes, respectively. The highest C_T_ where direct RT-LAMP detected SARS-CoV-2 RNA was 31.39 corresponding to a 1:40 dilution of a positive saliva sample with a starting C_T_ of 25.27. When RT-LAMP was performed on pools of SARS-CoV-2 negative saliva samples spiked with whole inactivated SARS-CoV-2 virus, RNA was detected at dilutions spanning 1:5 to 1:160 representing C_T_’s spanning 22.49-26.43. Here we describe a simple but critical rapid sample preparation method which can be used up front of RT-LAMP to permit direct detection of SARS-CoV-2 within saliva samples. Saliva is a sample which can be collected non-invasively without the use of highly skilled staff and critically can be obtained from both health care and home settings. Critically, this approach overcomes both the requirement and validation of different swabs and the global bottleneck observed in obtaining RNA extraction robots and reagents to enable molecular testing by PCR. Such testing opens the possibility of public health approaches for effective intervention to control the COVID-19 pandemic through regular SARS-CoV-2 testing at a population scale, combined with isolation and contact tracing for positive cases.

## Introduction

The COVID-19 pandemic caused by the SARS-CoV-2 virus poses a profound global threat to communities, economic activity and healthcare systems. It is generally accepted that a safe and efficacious vaccine will not be widely available in the immediate future whilst uncertainty remains over the trajectory of the pandemic. Moreover, herd immunity from a high proportion of the population having become immune to SARS-CoV-2 is not thought to be a viable public health strategy by most observers. One public health approach that has been advocated for suppression of the COVID-19 pandemic is regular SARS-CoV-2 testing at a population scale, combined with isolation and contact tracing for positive cases^1^. Such an approach requires a rapid relatively inexpensive diagnostic test for the presence of the SARS-CoV-2 virus, ideally based on samples that can be simply collected in both health care and non-health care (e.g. home) settings^2^.

The current international gold standard for diagnosis of infection with SARS-CoV-2 is detection of viral RNA by real-time Reverse Transcriptase Polymerase Chain Reaction (rRT-PCR) from a naso-pharyngeal or oropharyngeal swab in viral transport medium^3^. However, the procedure for collecting a good quality sample using this approach requires a degree of training and skill, potentially exposes the sampler to infectious droplets, and can be uncomfortable and traumatic for the patient, especially if undertaken frequently. Critically, supply issues during the pandemic have led to bottlenecks in the availability of reagents for molecular assays, leading to demand for bespoke extraction kits far outweighing available supply and hampering testing efforts globally. Alongside this, the requirement for swab testing has led to key manufacturers being unable to cope with swab demand for patient sampling^4,5^. This has meant that laboratories have had to undertake frequent and time-consuming assay validation on different swab types. As such, exploring alternative sample types and RNA detection methods that circumvent the issues above is an attractive solution.

Saliva is a sample which shows promise for infection diagnostics, including for diagnostic detection of coronaviruses and has been shown as a site where SARS-CoV-2 is found in early infection^6,7^. Collection of saliva is straightforward and can be done by the patient themselves using a drooling technique, and collection devices include a simple, widely available, universal plastic container.

Reverse-Transcription Loop-mediated isothermal AMPlification (RT-LAMP) is a highly sensitive reverse-transcription, autocycling, isothermal, strand displacement nucleic acid amplification technology^8^ which is more resistant to inhibitors than rRT-PCR), enabling simplification and even removal of the extraction procedure^9–11^. LAMP technologies have been applied for the detection of a wide range of pathogens^12–14^ including positive-sense RNA viruses^10^ and has been used extensively in the veterinary^15,16^ and plant industry^17–19^ and more recently as a human diagnostic^12,20,21^. At the height of the SARS-CoV-2 epidemic in the UK in early 2020, Hampshire Hospitals NHS Foundation Trust (HHFT) validated a novel RT-LAMP assay for the detection of SARS-CoV-2 RNA within nasopharyngeal and oropharyngeal swabs either directly from swab, or following RNA extraction^22^. For direct detection of SARS-CoV-2 RNA from swab, a simple dilution of 1:20 of the viral transport media in nuclease free water (NFW) was shown to be sufficient to overcome inhibition and to achieve sensitivity (DSe) and specificity (DSp) of 67% and 97%, respectively. When setting rRT-PCR cycle threshold (C_T_) cut-offs of ≤33 and ≤25, the DSe increased to 75% and 100%, respectively, with the specificity retained. Within this first study^22^, preliminary evaluation of Direct RT-LAMP for detection of SARS-CoV-2 in other clinical samples was performed using fourteen saliva samples collected from hospital in-patients confirmed from paired swabs as positive and negative for SARS-CoV-2. Using a 1:20 dilution of saliva in NFW, SARS-CoV-2 RNA was detected as expected in four of the positive swab samples but was unexpectedly detected in only two of the saliva samples. This indicated that more work was required to optimize the crude sample preparation method for detection of SARS-CoV-2 in saliva. Herein we describe the further optimisation of a simple sample preparation method to permit direct detection of SARS-CoV-2 within saliva samples using Direct RT-LAMP.

## Materials and Methods

### Virus isolates and clinical specimens

Optimisation of Direct RT-LAMP for detection of SARS-CoV-2 RNA in saliva was performed using three SARS-CoV-2 positive saliva samples collected from symptomatic patients at Hampshire Hospitals NHS Foundation Trust (HHFT) (n=1) and University Hospital Southampton (UHS) (n=2) who had previously had rRT-PCR positive SARS-CoV-2 positive naso-pharyngeal samples. An additional 15 SARS-CoV-2 negative saliva samples collected from asymptomatic UHS healthcare staff were used to prepare a pooled sample for specificity analysis. For spiking experiments, one pool of 25 SARS-CoV-2 negative saliva samples from asymptomatic UHS staff, and a second pool of 5 SARS-CoV-2 negative saliva samples also from asymptomatic UHS staff, were used to prepare pooled samples for spiking with whole inactivated virus (SARS-CoV-2 at ∼1×10^5^TCID_50_/ml was inactivated using beta-propriolactone (BPL). Collection of saliva involved the patient providing a fresh saliva sample into a 10ml universal container. Each positive saliva sample was diluted 1:1 in Mucolyse™ (active ingredient: dithiothreitol, Pro-Lab Diagnostics, UK) prior to dilution in either NFW or 10% Chelex^®^ 100 Resin (Bio-Rad Laboratories, Watford, UK)^23^. Mucolyse™ was also added 1:1 to the final pool of negative saliva samples and the SARS-CoV-2 spiked pools.

### RNA extraction

The saliva sample collected within the HHFT was extracted using the Maxwell^®^ RSC Viral Total Nucleic Acid Purification Kit (Promega UK Ltd., Southampton, UK) according to manufacturer’s instructions. Briefly, 200 µl of sample was added to 223 µl of prepared lysis solution (including 5 µl per reaction of Genesig^®^ Easy RNA Internal extraction control, Primerdesign Ltd, Chandler’s Ford, UK). Samples were then inactivated for 10 minutes at room temperature within the safety cabinet and 10 minutes at 56°C on a heat block before automated RNA extraction using a Maxwell^®^ RSC 48 Instrument (Promega UK Ltd., Southampton, UK). RNA was eluted in 50 µl of NFW.

The saliva samples collected from UHS were extracted using the MagMAX™CORE Nucleic acid purification kit (Thermofisher). Briefly, 10µl of sample (diluted in 190µl DEPC treated water) was added to 700µl of prepared lysis solution. Samples were then inactivated for 10 minutes at room temperature within the safety cabinet before automated RNA extraction using a KingfisherFlex (Thermofisher). RNA was eluted in 90 µl of NFW.

### Real-time reverse-transcription PCR (rRT-PCR)

All rRT-PCR assays were performed in single replicates using 5 µl of RNA template. The saliva sample collected within the HHFT was analysed using the COVID-19 genesig^®^ Real-Time PCR assay (Primerdesign Ltd, Chandler’s Ford, UK) according to the manufacturer’s guidelines, on a MIC qPCR Cycler (Bio Molecular Systems, London, UK). The cycling conditions were adjusted to the following: a reverse-transcription (RT) step of 10 minutes at 55°C, a hot-start step of 2 minutes at 95°C, and then 45 cycles of 95°C for 10 seconds and 60°C for 30 seconds. The Genesig^®^ COVID-19 positive control included in the kit, a negative extraction control, and a no template control were also included on each rRT-PCR run.

The saliva samples collected from the UHS and the spiked whole virus dilution series were tested using the E gene RT-PCR as described previously (Corman et al.,2020) using the AgPath-ID™ PCR kit (Thermofisher). Samples were run on an Aria qPCR Cycler (Agilent) and results analysed using the Agilent AriaMX 1.5 software. The cycling conditions were adjusted to the following: a reverse-transcription (RT) step of 10 minutes at 55X°C, a hot-start step of 3 minutes at 94°C, and then 45 cycles of 94°C for 15 seconds and 60°C for 15-30 seconds during data acquisition. The SARS-CoV2 positive control RNA, a negative extraction control, and a no template control were also included on each rRT-PCR run.

### Preparation of 10% (w/v) Chelex^®^ 100 Resin

10% (w/v) Chelex^®^ 100 Resin was made up by resuspending Chelex^®^ 100 Resin (200-400 mesh) (Bio-Rad Laboratories, catalogue number #142-1253) in Milli-Q^®^ water at 10% (w/v). The solution was heated at 70°C for 30 minutes. Two washes in Milli-Q^®^ water were performed by allowing the Chelex^®^ 100 Resin to settle, removing the supernatant, adding Milli-Q^®^ water to 10% (w/v) and shaking. After a second wash, Milli-Q^®^ water was added to give a final 10% Chelex^®^ 100 (w/v) Resin solution.

### Reverse-transcription loop-mediated isothermal amplification (RT-LAMP)

RT-LAMP reactions were performed using OptiGene Ltd. (Camberley, UK) COVID-19_Direct RT-LAMP KIT-500 kit which targets the *ORF1ab* region of the SARS-CoV-2 genome.

Each RT-LAMP reaction consisted of: 17.5 μl of RT-LAMP Isothermal Mastermix (containing 8 units of GspSSD2.0 DNA Polymerase, 7.5 units of Opti-RT reverse transcriptase and a proprietary fluorescent dsDNA intercalating dye and a proprietary enhancing enzyme), 2.5 μl of 10X COVID-19 Primer Mix, and 5 μl of RNA/sample. RT-LAMP reactions were performed in duplicate at 65°C for 20 mins on a Genie^®^ HT (OptiGene Ltd., UK). An exponential increase in fluorescence (ΔF) indicated a positive reaction, which was quantified by a time to positivity (Tp) value, called at the point where the fluorescence level on the amplification curve, crosses the threshold of 5000. To confirm the specificity of the amplification reaction, an anneal curve was performed: RT-LAMP products were heated to 98°C for 1 min, then cooled to 80°C decreasing the temperature by 0.05°C/s.

Genie^®^ embedded software (OptiGene Ltd., UK) was utilised to analyse RT-LAMP results and define thresholds for result calling. All RT-LAMP reactions were performed in duplicate, and a sample was considered positive when a Tp was observed in at least one replicate with amplification above 5000 fluorescence points and had an anneal temperature of between 81.50°C and 84.05°C with a derivative above 2500 F/°C.

For Direct RT-LAMP, 5 μl of saliva diluted in NFW or 10% (w/v) Chelex^®^ 100 Resin (Bio-rad) spanning 1 in 5 to 1 in 640, with and without heat treatment (70°C for 4 minutes or 98°C for 2 mins) was added to the reaction. Heating was performed on a dry heat block. The same treatments were applied to the saliva pools spiked with whole inactivated virus and to the non-spiked negative saliva pool, however the first spiked saliva pool was only titrated as far as 1:40 in the first instance. After addition to direct RT-LAMP all treatments were pooled according to dilution (e.g. all temperature treatments were pooled according to the dilution) and extracted for rRT-PCR analysis.

## Results

### Optimisation of the Direct-RT-LAMP

Optimization of the Direct RT-LAMP assay for detection of SARS-CoV-2 was determined using three positive saliva samples, a pool of non-spiked negative saliva and a pool of spiked saliva.

The three positive saliva samples diluted 1:1 in Mucolyse™ rRT-PCR C_T_ values were 21.08, 24.47 and 25.27 (Table 1). For the first spiked saliva pool, the whole inactivated virus spiked into saliva prior to dilution rRT-PCR C_T_ was 26.70 (Table 2). For the second spiked saliva pool, the whole inactivated virus spiked into saliva diluted 1:1 in Mucolyse™ (prior to further dilutions) rRT-PCR C_T_ was 22.86 (Table 3).

**Table 1.**
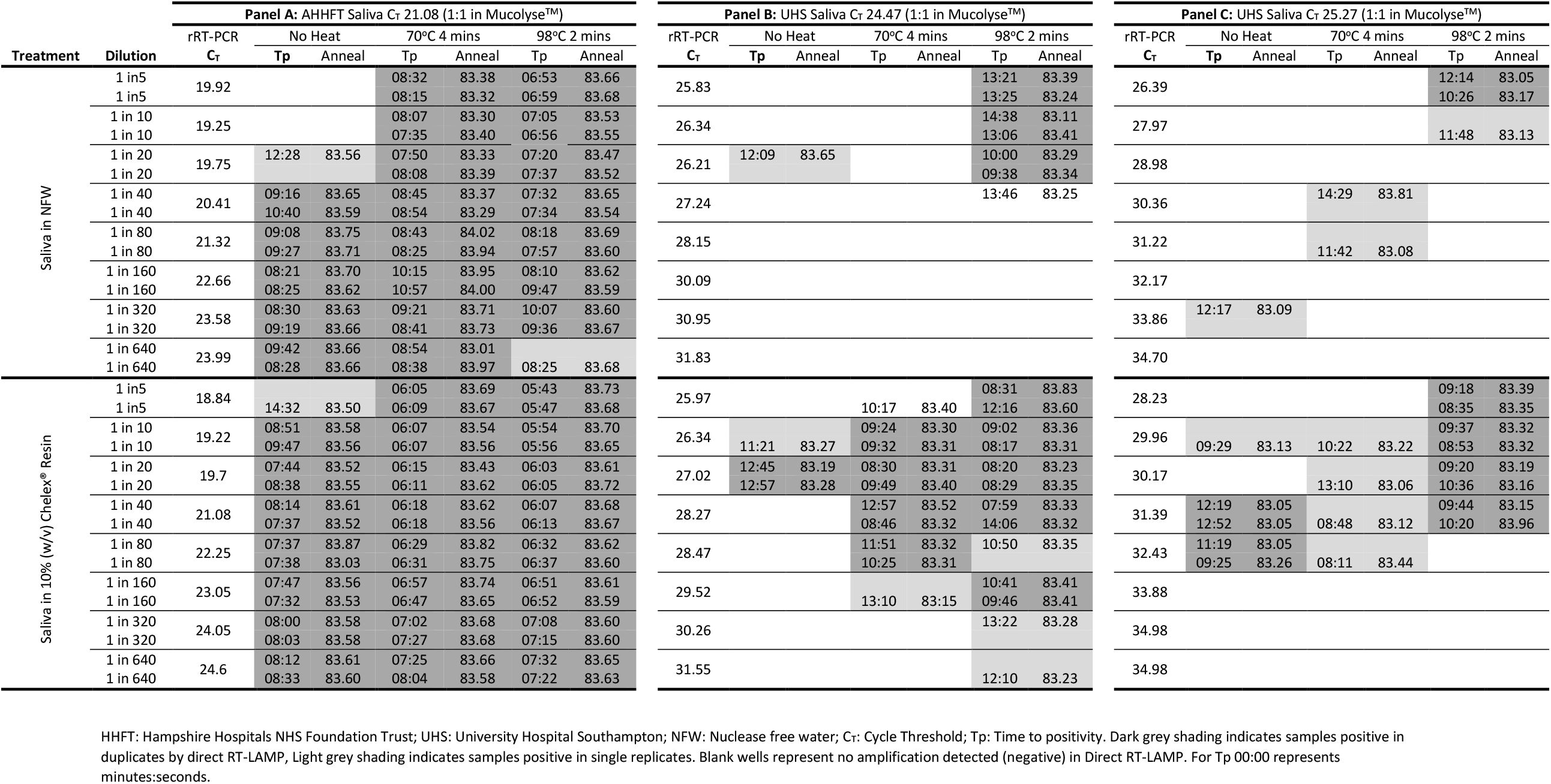
Sample preparation optimisation for direct detection of SARS-CoV-2 in crude saliva

**Table 2:**
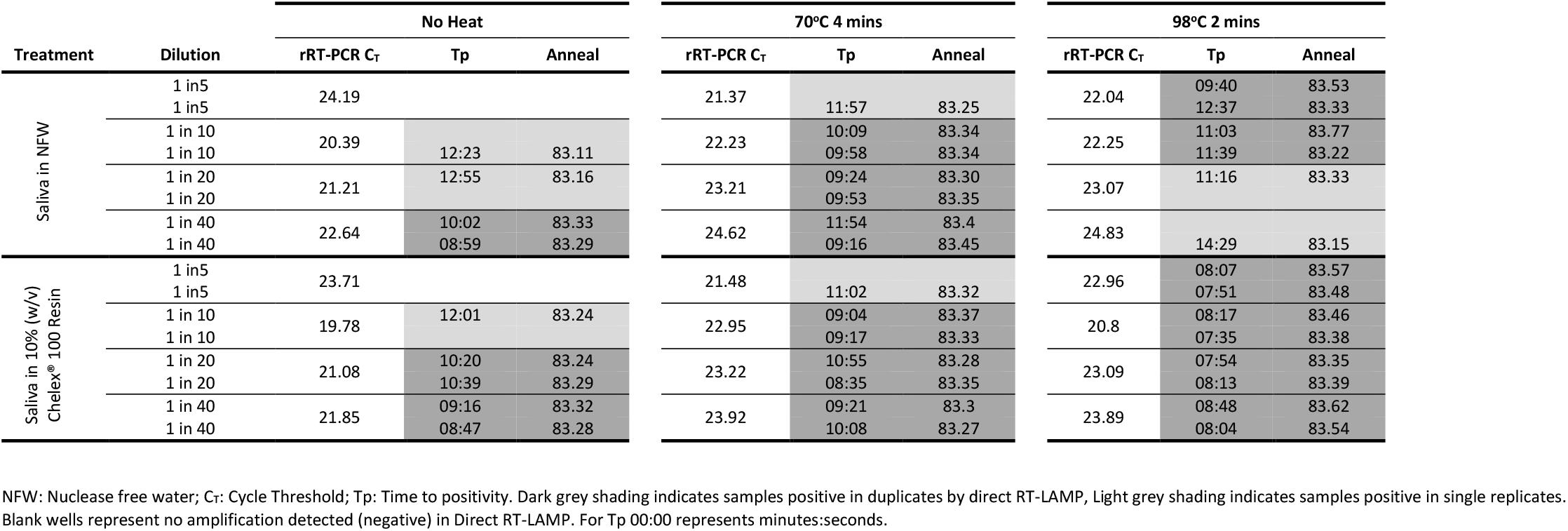
Dilution series of inactivated whole virus spiked into pooled saliva from 25 negative patient samples (no predilution 1:1 in Mucolyse™)

**Table 3:**
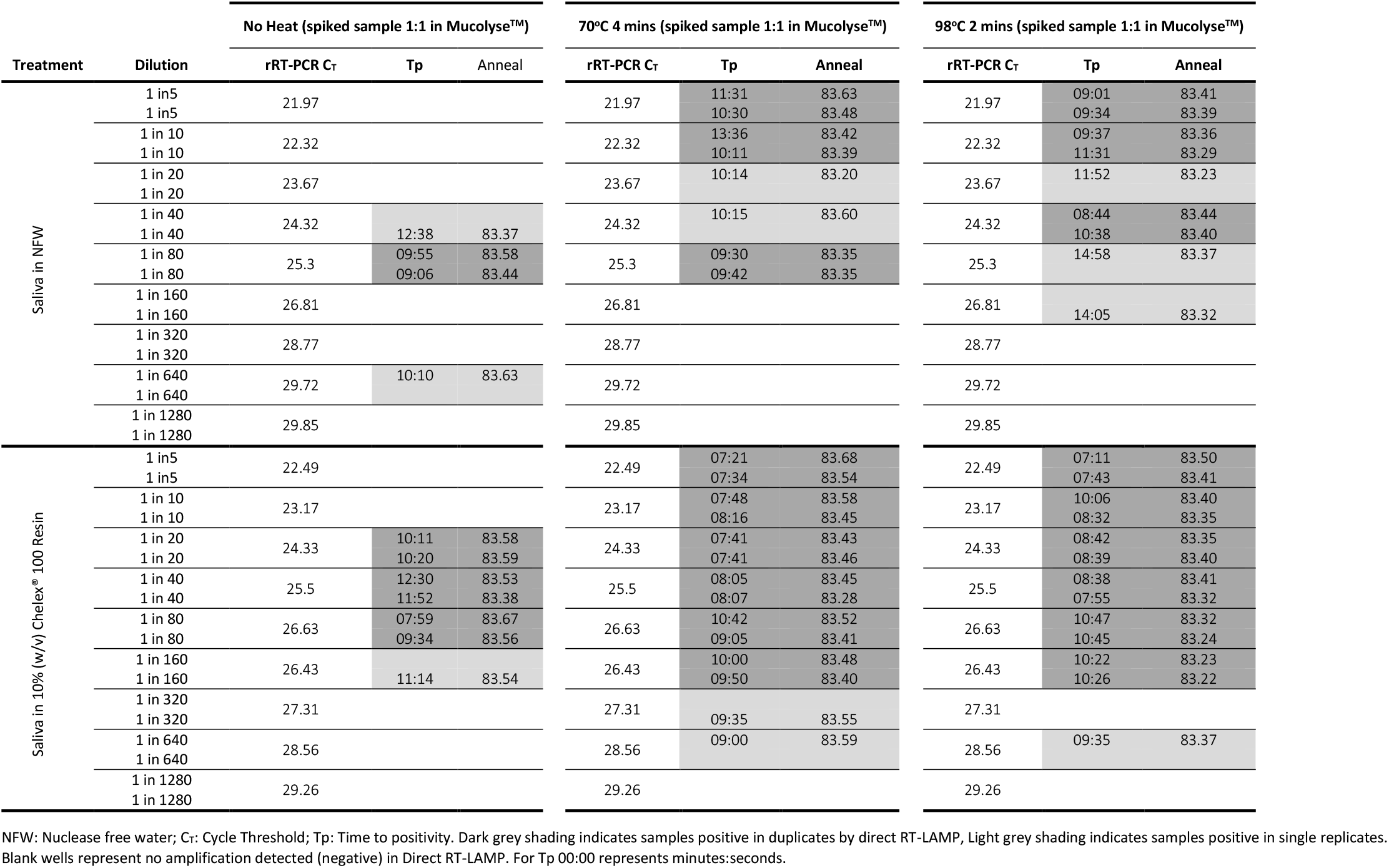
Dilution series of inactivated whole virus spiked into pooled saliva from five negative patient samples (predilution 1:1 in Mucolyse™)

From rRT-PCR data samples were assessed for sensitivity using the Direct-LAMP protocol. Samples were assessed in order of highest viral load by rRT-PCR (C_T_ 21.08: Table 1, Panel A) result to lowest (C_T_ 25.27: Table 1, Panel C).

The saliva sample with the highest viral load (C_T_ 21.08) when diluted in water was detected in duplicate in five dilutions (1:40 to 1:640) without heat treatment, in all eight dilutions (1:5 to 1:640) following 70°C for 4 mins and in seven dilutions 1 in 5 to 1 in 640 following 98°C for 2 mins (Table 1, Panel A). When diluted in 10% (w/v) Chelex^®^ 100 Resin the same saliva sample (C_T_ 21.08) was detected in duplicate in seven dilutions (1:10 to 1:640) without heat treatment and in all eight dilutions (1:5 to 1:640) following either 70°C for 4 minutes or 98oC for 2 minutes (Table 1, Panel A).

The saliva sample with a C_T_ of 24.47 when diluted in water was not detected in duplicate in any dilution without heat or following 70°C for 4 mins (Table 1, Panel B). This sample was detected in duplicate in three dilutions (1:5 to 1 in 20) only following 98°C for 2 mins (table 1). When diluted in 10% (w/v) Chelex^®^ 100 Resin the same saliva sample (C_T_ 24.47) was detected in duplicate in one dilution (1:20) without heat treatment, in 4 dilutions (1:10 to 1:80) following 70°C for 4 minutes and in five dilutions (1:5 to 1:40 and 1: 160) following 98°C for 2 minutes (Table 1, Panel B).

The saliva sample with the lowest viral load (C_T_ 25.27) when diluted in water was not detected in duplicate in any dilution without heat or following 70°C for 4 mins (Table 1, Panel C). This sample was detected in duplicate in one dilution (1:5) only following 98°C for 4 mins (Table 1, Panel C). When diluted in 10% (w/v) Chelex^®^ 100 Resin the same saliva sample (C_T_ 25.27) was detected in duplicate in two dilutions (1:40 and 1:80) without heat treatment, in no dilutions following 70°C for 4 minutes and in four dilutions (1:5 to 1:40) following 98°C for 2 minutes (Table 1, Panel C).

The pool of saliva samples negative for SARS-CoV-2 was negative also on Direct RT-LAMP for all assay conditions (data not shown as all samples reported a negative result).

The whole inactivate virus spiked into saliva with a C_T_ of 26.70 when diluted in water was detected in duplicate at one dilution (1:40) without heat, in three dilutions (1:10, 1:20, 1:40) following 70°C for 4 mins and in two dilutions (1:5 and 1:10) following 98°C for 2 mins (Table 2). When diluted in 10% (w/v) Chelex^®^ 100 Resin the same saliva sample was detected in duplicate in two dilutions (1:20 and 1:40) without heat treatment, in three dilutions (1:10, 1:20, 1:40) following 70°C for 4 minutes and in all four dilutions (1:5 to 1:40) following 98°C for 2 minutes (Table 2).

A further dilution series of SARS-CoV-2 inactivated whole virus was prepared to include the 1:1 Mucolyse™ dilution that is used for clinical samples and to extend beyond a 1:40 dilution to reach the limit of detection of the Direct RT-LAMP assay. The whole inactivated virus spiked into saliva (C_T_ of 22.86 when diluted in water was detected in duplicate at one dilution (1:80) without heat, in three dilutions (1:5, 1:10 and 1:80) following 70°C for 4 mins and in three dilutions (1:5, 1:10 and 1:40) following 98°C for 2 mins (Table 3). When diluted in 10% (w/v) Chelex^®^ 100 Resin the same saliva sample was detected in duplicate in three dilutions (1:20, 1:40 and 1:80) without heat treatment, in six dilutions (1:5, 1:10, 1:20, 1:40, 1:80 and 1:160) following 70°C for 4 minutes and in six dilutions (1:5, 1:10, 1:20, 1:40, 1:80, 1:160) following 98°C for 2 minutes (Table 3).

## Discussion

This study describes the rapid optimization of a method to permit direct detection of SARS-CoV-2 RNA within saliva samples using RT-LAMP, without need for prior RNA extraction. Our previous publication was focused on optimizing conditions for rapid detection of SARS-CoV-2 within viral transport media from swabs samples^22^. In that publication, preliminary evaluation of the direct transfer of the swab sample preparation method for comparable detection in paired saliva samples was poor, indicating that a different sample preparation method would be required for optimal detection of SARS-CoV-2 RNA in crude saliva. In this study we show for the first time that the optimal sample preparation method to allow SARS-CoV-2 RNA detection within crude saliva samples (1:1 mix of saliva and Mucolyse™ (active ingredient dithiothreitol)) requires saliva dilution in 10% (w/v) Chelex^®^ 100 Resin and heating to 98°C 2 minutes prior to adding to the direct RT-LAMP reagents.

When using this approach SARS-CoV-2 RNA was reliably detected in duplicates for a wide range of dilutions assessed from positive saliva samples with a starting C_T_ value of 21.08, 24.47 and 25.27. The combination of a chelating resin (Chelex^®^ 100 Resin) and heating the sample to 98°C successfully overcame matrix inhibition and or matrix “protection” of viral capsid nucleic acid release which was observed in the samples which did not receive this protocol. The time to positivity (speed at which SARS-CoV-2 RNA was detected in saliva) was proportional to the “strength” of the saliva sample after addition of 1:1 Mucolyse™ (active ingredient: dithiothreitol) as determined by real-time reverse-transcriptase PCR with SARS-CoV-2 detected in 05:55 (C_T_ 21.08), 08:39 (C_T_ 24.47) and 09:15 (C_T_ 25.27) minutes, respectively when using a dilution of 1:10 of saliva into 10% (w/v) Chelex^®^ 100 Resin. Importantly, using this method, no amplification was detected in the negative pooled saliva samples, confirming the compatibility of this sample preparation approach in maintaining specificity of the assay.

Due to the lower prevalence of SARS-CoV-2 at the time of optimisation of this sample preparation method, only three positive saliva samples were available for analysis. To strengthen conclusions drawn from these clinical specimens spiked dilution series of whole inactivated virus spiked into pooled saliva were also evaluated with equivalent results obtained. This method should therefore be translatable to saliva samples regardless of whether they are obtained from symptomatic or asymptomatic patients.

Studies in macaque monkeys demonstrated that the salivary glands in the mouth are the first site in the body to be affected by SARS-CoV infection^24^ and several groups have reported high sensitivity and specificity saliva of rRT-PCR for SARS-CoV-2 in COVID-19 patients^25,26^. SARS-CoV-2 is therefore present in saliva samples early in the course of infection and can be spread to other individuals efficiently through salivary droplets generated when talking loudly or singing^27^. Population screening of saliva samples may therefore be an effective strategy to detect the important group of people who are infectious but not yet symptomatic. There is also evidence that SARS-CoV-2 may be present in saliva during the recovery phase from infection after upper respiratory samples have become negative^28^, making saliva an attractive sample type for identification of individuals in the population who could transmit infection^4^.

These findings add to the increasing literature supporting saliva as a reliable sample type in which to detect SARS-Cov-2 RNA. Using saliva samples collected in a simple collection pot, we described an approach that paves the way for a rapid diagnostic test for the presence of the SARS-CoV-2 virus based on samples that can simply be collected at home or in other non-health care settings. Critically, this approach overcomes both the requirement and validation of different swabs and the global bottleneck observed in obtaining RNA extraction robots and reagents to enable molecular testing by PCR. Such testing opens the possibility of public health approaches for suppression of the COVID-19 pandemic through regular SARS-CoV-2 testing at a population scale at relatively low cost, combined with isolation and contact tracing for positive cases.

## Data Availability

All data is contained within the tables in the manuscript

## Ethics

UHS saliva collection and analysis was conducted with informed written consent following institutional review board approval (ENACT – Enabling New Approaches for CoVID-19 Treatment).

